# Experimental human pneumococcal carriage in adults with HIV in Malawi

**DOI:** 10.64898/2026.05.13.26353107

**Authors:** K Doherty, AE Chirwa, E Nsomba, V Nkhoma, B Galafa, G Kadzanja, M Mailboy, E Mangtani, S Songolo, G Lipunga, A Sigoloti, T Harawa, C Mkandawire, MP Kamanga, N Toto, L Makhaza, J Ndaferankhande, AR Noel, M Al-Habbal, S Mbewe, T Nthandira, L Chimgoneko, G Tembo, J Phiri, J Reiné, T Chikaonda, MYR Henrion, DM Ferreira, H Mwandumba, NPK Banda, K Jambo, SB Gordon, the MARVELS consortium

## Abstract

**Background:** People living with HIV (PLHIV) in sub-Saharan Africa exhibit high rates of pneumococcal carriage compared to HIV-uninfected adults, despite antiretroviral therapy. We established a novel controlled human infection model of experimental pneumococcal carriage in people living with HIV to understand carriage dynamics in this at-risk population.

**Methods:** Seventy-five virally suppressed and clinically stable PLHIV and 75 HIV-uninfected controls were inoculated with escalating doses of pneumococcus serotype 6B. Carriage acquisition and density were determined by microbiological culture of nasal wash samples collected before and up to 14 days after inoculation. Adverse events were identified by active and passive surveillance. Participant-reported acceptability was established using a Likert scale.

**Findings:** No serious adverse events occurred. Mild adverse events were similar between groups (19% [14/75] in PLHIV, 13% [10/75] in HIV-uninfected; p=0·505). More than 90% of participants reported acceptability with all study procedures. Experimental carriage occurred in 21% (16/75) of PLHIV compared with 36% (27/75) of HIV-uninfected participants (adjusted odds ratio 0·39 [95% CI 0·16-0·91]). Among PLHIV without detectable cotrimoxazole, 28% (8/29) acquired experimental carriage. Carriage clearance rates were lower in PLHIV (hazard ratio 0·44 [95% CI 0·14-1·42]).

**Interpretation:** In carefully selected PLHIV with effective viral suppression and clinical stability experimental pneumococcal carriage acquisition did not exceed that in HIV-uninfected adults, even after accounting for antibiotic use, natural pneumococcal co-colonisation, and sociodemographic differences. These findings suggest that high carriage prevalence in PLHIV in sub-Saharan Africa may be driven more by prolonged carriage duration than increased susceptibility to acquisition. This model provides a platform to investigate mechanisms underlying carriage susceptibility and impaired clearance in PLHIV and to evaluate interventions aimed at reducing the carriage burden in sub-Saharan Africa.

**Funding:** Wellcome Trust

## Introduction

People living with HIV (PLHIV) in sub-Saharan Africa experience a burden of pneumococcal disease up to 30 times that of HIV-uninfected counterparts and exhibit pneumococcal carriage rates 50% higher, even when established on antiretroviral therapy (ART).^1,2^ PLHIV in Malawi exhibit higher rate, density, and duration of carriage, and shed more pneumococcus than their HIV-uninfected counterparts.^1,3^ The high carriage burden in PLHIV is one driver of the high force of infection which is overcoming vaccine-induced herd immunity efforts in Malawi and is a driver antimicrobial resistance in the country.^3–5^ Despite this, there is currently no pneumococcal vaccination strategy for addressing the burden of pneumococcal carriage and disease among PLHIV in sub-Saharan Africa. The pneumococcal polysaccharide vaccine (PPV23) was associated with excess pneumococcal disease in PLHIV in Uganda compared to placebo, limiting its use in this context.^6^ The first generation pneumococcal conjugate vaccine (PCV7) was safe and effective as secondary prophylaxis against vaccine-serotype pneumococcal disease in Malawi.^7^ No study has evaluated the newer generation PCVs nor vaccine efficacy against carriage in adult PLHIV. PLHIV constitute 8·9% of the adult community in Malawi and 14% in an urban setting, making such data key to guide policy makers on the potential for herd immunity and to identify the most effective vaccination strategy for PLHIV.^8,9^

Controlled human infection models (CHIMs) can accelerate vaccine evaluations and development and can further our understanding of susceptibility to infectious diseases.^10^ They are more cost-effective than large-scale community trials. To date most CHIMs have been conducted in young, healthy adults in non-endemic regions who are least affected by infectious diseases. More recently established CHIMs focus on specific populations with the greatest infectious disease burden.^11,12^ We describe results from an Experimental Human Pneumococcal Carriage (EHPC) model in PLHIV in Malawi which is the first CHIM in this important population. We aimed to demonstrate the safety and acceptability of EHPC in PLHIV in Malawi and describe experimental pneumococcal carriage dynamics in virally suppressed and clinically stable PLHIV compared to HIV-uninfected controls.

## Materials and methods

### Study design

Successive cohorts of eligible participants were inoculated intranasally with a known colony-forming unit dose of live, penicillin-sensitive, *Streptococcus pneumoniae* serotype 6B at successively increasing doses. Participants attended clinic on days 2, 7, and 13 or 14 post-inoculation to detect experimental pneumococcal carriage and for proactive adverse event surveillance, as described in the published study protocol and Supplementary Table E1.^13^ In a second phase of the study, participants who remained carriage-negative underwent repeat inoculation on day 14 to increase likelihood of experimental carriage (Supplementary Figure E1). Double-inoculated participants were followed up on days 16, 21, and 28. Ethical approval was granted by the Malawian National Health Science Research Committee and by Liverpool School of Tropical Medicine’s Research Ethics Committee. The trial was prospectively registered with ClinicalTrials.gov (NCT05698225). This study is reported in accordance with Consolidated Standards of Reporting Trials guidelines (Supplementary Table E2).^14^

### Study population and setting

Participants aged 25–45 years with HIV and HIV-uninfected controls were recruited between June 2023 and August 2024 through district health clinics and institutional research registers. Eligibility criteria are detailed in the published protocol.^13^ In brief, PLHIV deemed safe for experimental inoculation had to demonstrate evidence of antiretroviral therapy for at least two years and absence of comorbidities or severe illness in preceding two years. Screening tests confirmed HIV viral suppression and a CD4 count >350 cells/mm^3^ prior to inoculation. Participants with a household contact at increased risk of pneumococcal disease such as children <5 years were excluded. Participants were excluded if they had had a respiratory tract infection or antibiotic course in the preceding two weeks (aside from prophylactic cotrimoxazole used by PLHIV in Malawi). Participants with prior receipt of any pneumococcal vaccine or natural pneumococcal serotype 6B carriage at baseline by microbiological culture were excluded (carriers of any other serotype were eligible for participation).

### Pneumococcal challenge and safety monitoring

The serotype 6B challenge strain (BHN418) was prepared as previously described.^15^ A pre-specified dose of bacterium was instilled into each nostril. Doses were escalated from 20000 to 320000 colony-forming units per nostril according to predefined safety and carriage criteria.^13^ Participants were observed in open study accommodation for three days post-inoculation and monitored thereafter through scheduled visits and telephone contact. Adverse events were actively solicited and vital signs recorded at each visit. Serious adverse events were defined as death, hospitalisation, life-threatening events, or permanent disability. An independent Data and Safety Monitoring Board reviewed data monthly. Carriage-positive participants received amoxicillin at study completion. Acceptability was assessed using a structured exit questionnaire (Supplementary Table E3).

### Outcome measure: Experimental pneumococcal carriage

Experimental pneumococcal carriage was defined as detection of serotype 6B in nasal wash samples at any post-inoculation time-point by microbiological culture. Full laboratory procedures are described in Supplementary Materials.

### Other variables

Demographic and clinical data were collected by questionnaire and medical record review. Serum cotrimoxazole concentrations were measured and any detectable level was considered evidence of prophylaxis.

### Sample size and statistical analysis

The sample size of 150 was estimated to detect a difference in carriage with 80% power and two-sided significance level of 0·05 using community estimates of carriage prevalence.^13^ Statistical analyses are described in Supplementary Materials. All analyses were performed in R version 4.3.0.^13^

## Results

### Comparison groups and sociodemographic characteristics

Seventy-five PLHIV and 75 HIV-uninfected controls were inoculated with pneumococcus serotype 6B at progressively increasing doses and included in analysis (Supplementary Figure E1 for study population creation and dose escalation). The demographic details of analysed participants are shown in Table 1. The PLHIV group were older than HIV-uninfected (median age 38·0 years (interquartile range [IQR] 35·0-42·0) versus 32·0 (IQR 28·5-36·0). The PLHIV group were majority female compared to a more sex-balanced HIV-uninfected group (57/75 [76%] female PLHIV; 39/75 [52%] female HIV-uninfected). Both groups had a similar rate of natural pneumococcal carriage with 33% (25/75) of PLHIV and 31% (23/75) of HIV-uninfected displaying natural pneumococcal carriage at baseline of both PCV13 vaccine and non-vaccine serotypes (Figure 1 and Supplementary Table E4).

**Table 1.** Baseline characteristics.

|  | **PLHIV**<br>**n=75** | **HIV-uninfected**<br>**n=75** |
| --- | --- | --- |
| *Female sex* | 76% (n=57) | 52% (n=39) |
| *Age (median, IQR)* | 38.0 (35.0 - 42.0) | 32.0 (28.5 - 36.0) |
| *Natural non-6B carriage at baseline* | 33% (n=25/75) | 31% (n=23/75) |
| *PCV13 serotype* | 13% (n=10/75) | 8.0% (n=6/75) |
| *Non-PCV13 serotype* | 20% (n=15/75) | 23% (n=17/75) |
| *Baseline CD4 count (median, IQR)* | 622 (473-807) | .. |
| *Detectable viral load (n)* | 0 | .. |
| *Years since HIV diagnosis (median, IQR)* | 9.3 (5.8-13.0) | .. |
| *Years since initiation of ART (median, IQR)* | 8.2 (5.5-12.4) | .. |
| *Lag between HIV diagnosis and ART initiation (median, IQR)* | 0.0 (0.0-0.0) | .. |
| *Current ART regime* |  |  |
| *TDF/3TC/DTG* | 97% (n=73) | .. |
| *ABC/3TC/DTG* | 1.3% (n=1) | .. |
| *TDF/DTG/EFV* | 1.3% (n=1) | .. |
| *Self-reported cotrimoxazole use* | 72% (n=54) | .. |

*IQR=interquartile range, PCV13=13-valent pneumococcal conjugate vaccine, ART=antiretroviral therapy, TDF=tenofovir disoproxil, 3TC=levetiracetam, DTG=dolutegravir, ABC=abacavir, EFV=efavirenz*

**Figure 1.**
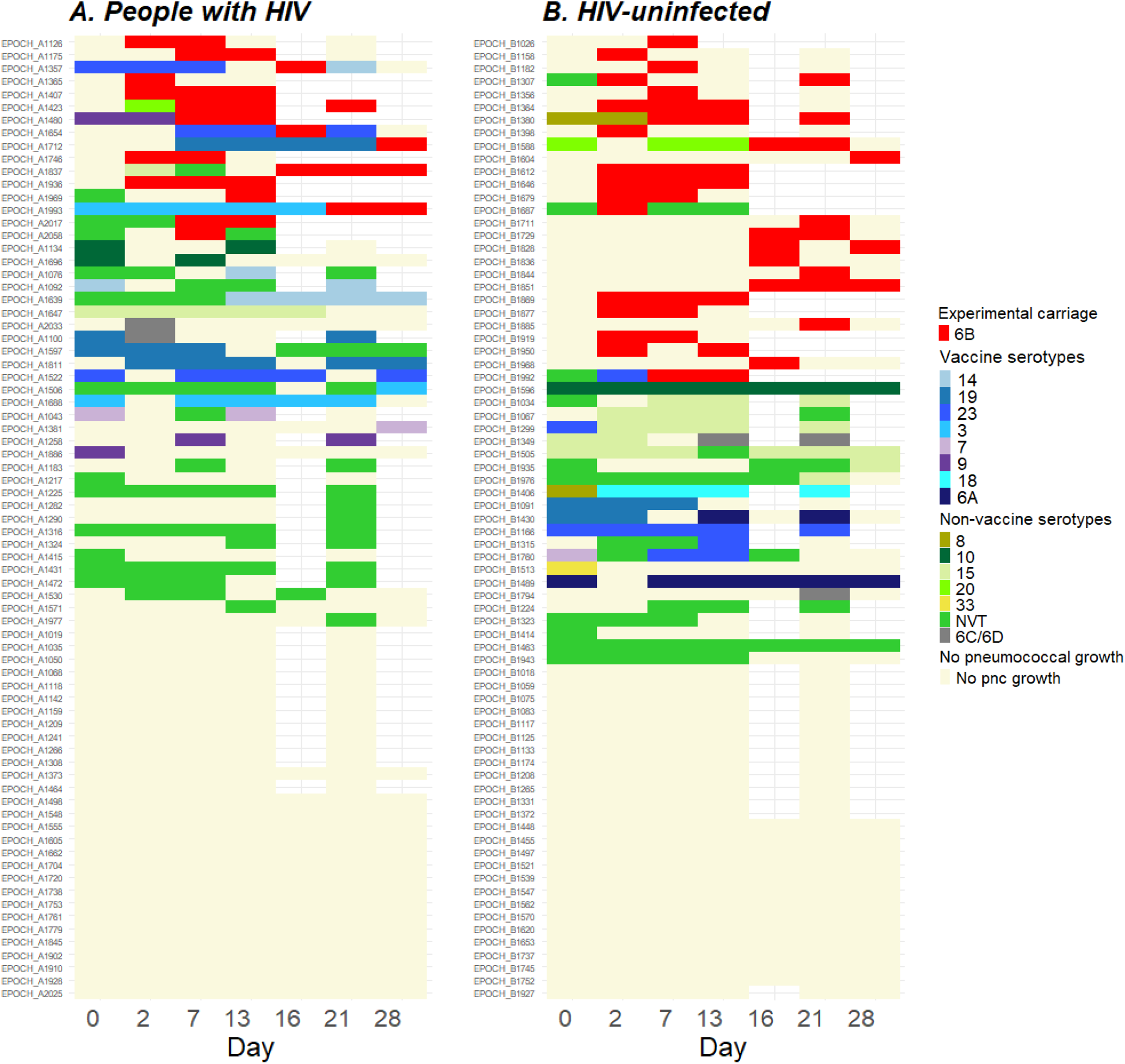
Experimental and natural carriage at baseline and following experimental pneumococcal challenge. Experimental (red), vaccine serotype natural (blue-purple shades), and non-vaccine serotype natural (green-yellow shades) pneumococcal carriage at baseline (day 0) and following experimental pneumococcal inoculation (days 2 to 28) identified by classical culture and latex agglutination of nasal wash samples. Each row represents an individual participant. Blank cells reflect no sample taken on that day. Vaccine serotypes are pneumococcal serotypes which PCV13 protects from, and non-vaccine serotypes are all other pneumococcal serotypes. pnc= pneumococcal

As specified in the inclusion criteria, all PLHIV had a CD4 T-cell count over 350 cells/mm^3^ (median 622 [IQR 473-807]) and an undetectable viral load at baseline. The median time since HIV diagnosis was 9·3 years (IQR 5·8-13·0 years), and most participants had initiated antiretroviral therapy (ART) as soon as they were diagnosed. Ninety-seven percent (73/75) of PLHIV were on the country’s first-line ART regime (Tenofovir/Levetiracetam/Dolutegravir).

### Experimental pneumococcal serotype 6B carriage is safe and acceptable in people living with HIV in Malawi

There were no serious adverse events in this study. No statistically significant difference in the rate of mild adverse events was detected between PLHIV and HIV-uninfected (18·7% [14/75] in PLHIV, 13·3% [10/75] in HIV-uninfected controls, p=0·505) (Figure 2A). Mild adverse events included symptoms possibly related to inoculation (e.g. sore throat, cough, coryzal symptoms), and other symptoms unrelated to inoculation (e.g. abdominal pain, diarrhoea, vomiting) (Supplementary Table E5). One PLHIV participant required additional study visits however this was unrelated to the inoculation (diarrhoeal illness commencing before inoculation with full recovery). No difference in adverse event rate was detected between participants with and without experimental pneumococcal carriage (Supplementary Table E6).

**Figure 2.**
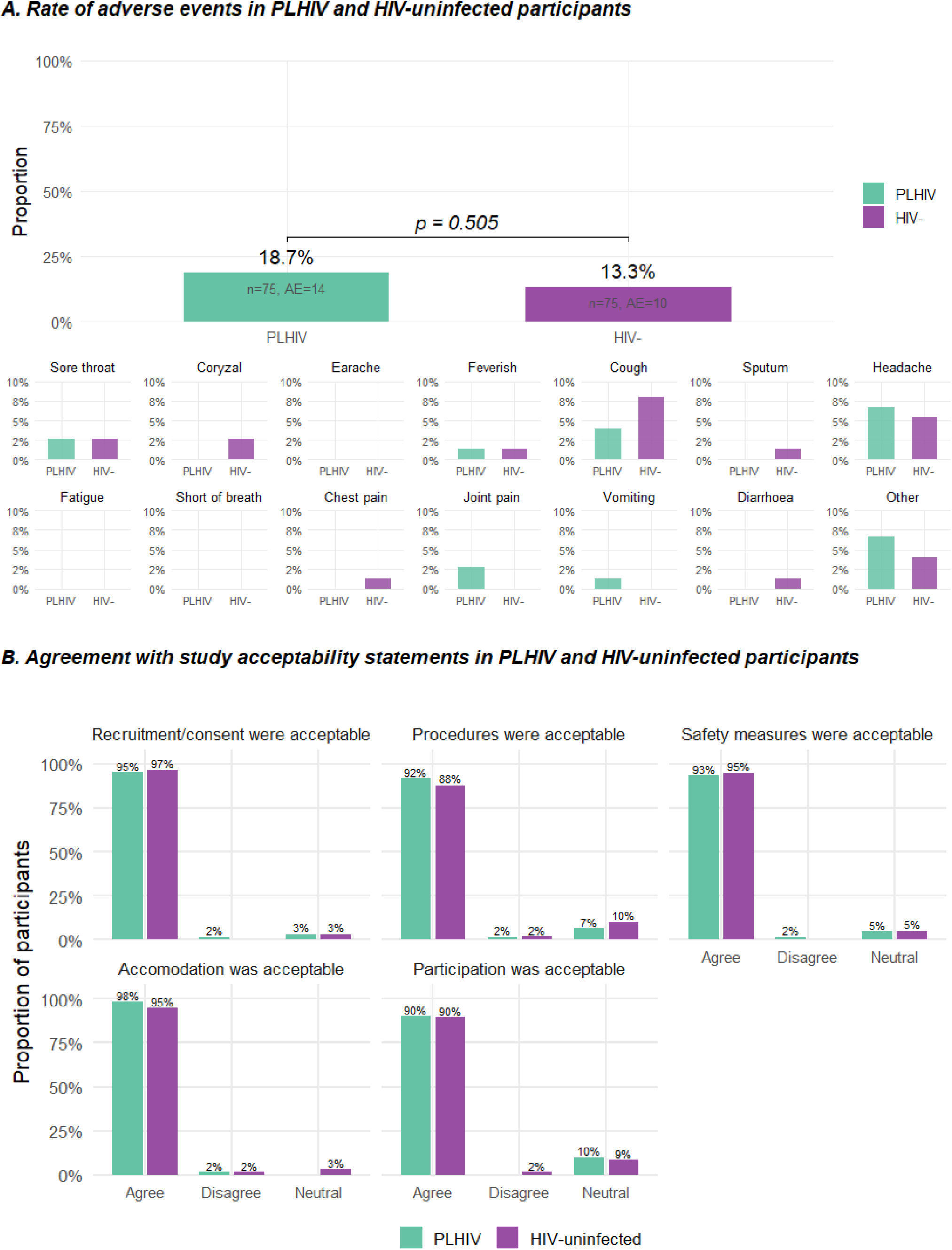
Safety and acceptability of experimental pneumococcal carriage in PLHIV and HIV-uninfected adults. (A) Proportion of PLHIV (turquoise) and HIV-uninfected (purple) who described an adverse event following direct solicitation during one of the study follow-up visits (days 2, 7, 13/14, 16, 21, or 28). Proportion=number of participants with an adverse event at any follow-up visit/total number of participants in each group. P-value represents likelihood results are observed by chance using a two-sided Wilcoxon signed-rank test. The specific symptom bar plots represent the symptoms which were specifically solicited from participants at each study visits. Other symptoms included abdominal pain (n=3 PLHIV), itchy eyelid (n=1 PLHIV), rash (n=1 HIV-uninfected), and unknown (n=1 PLHIV). All adverse events described in the box plots were deemed to be mild by a study clinican and none required additional study visits. (B) Acceptability of study to PLHIV (turquoise) and HIV-uninfected (purple) participants at study-end measured on a Likert-scale. Each boxplot represents the lowest scoring answer to multiple statements related to each of the five domains. The statements for each domain were as follows: 1. Recruitment and consent: “The way I heard about this study was appropriate and did not interfere with my clinical care”, “I received sufficient information about the study before consenting to participate”, and “I was given sufficient time to consider the study before consenting to participate”; 2. Study procedures: “The procedure to have the bacteria placed in my nose did not cause undue discomfort” and “The procedures to take samples from my nose did not cause undue discomfort”; 3. Study design and safety: “The frequency of visits during the study was appropriate”, “I felt the steps taken to keep me safe during the study were appropriate (for example; daily text messages with study team, health-checks during study visits, study accommodation)”, and “I was able to contact the study team easily for advice”; 4. Study accommodation: “The accommodation provided after inoculation was satisfactory” and “I felt 3 days was an appropriate time to remain in study accommodation”; 5. Study participation: “The clinical team treated me with respect and kindness throughout the study”, “The compensation provided by the study was appropriate”, and “I would recommend participation in this study to a friend”. Participants could answer 5=strongly agree, 4=agree, 3=neutral, 2=disagree, and 1=strongly disagree with strongly agree/agree and strongly disagree/disagree combined into one bar on the boxplot. PLHIV= people living with HIV HIV- = HIV-uninfected adults.

Acceptability was high and did not differ between PLHIV and HIV-uninfected in relation to the five measured study domains (Figure 2B). Over 97% of participants agreed that recruitment and consent procedures, study safety and follow-up procedures, and study accommodation were appropriate and sufficient (Supplementary Table E3). Among both PLHIV and HIV-uninfected participants 99% would recommend participation in the study.

### Experimental pneumococcal serotype 6B carriage rate unexpectedly lower in people living with HIV than in HIV-uninfected

In total, 21% of PLHIV (16/75, 95% confidence interval [95% CI] 13%-32%) and 36% of HIV-uninfected participants (27/75, 95% CI 25%-48%) experienced experimental pneumococcal carriage (defined as the presence of live serotype 6B pneumococcus in cultured nasal wash samples at any time-point following inoculation) (Table 2 and Figure 3A). Eleven PLHIV (11/75, 15%) experienced experimental carriage after first inoculation and an additional five (5/37, 14%) after second inoculation. Seventeen HIV-uninfected (17/75, 23%) experienced experimental carriage after first inoculation and an additional ten (10/35, 29%) after second inoculation. The odds ratio (OR) of experimental pneumococcal carriage in PLHIV compared to HIV-uninfected was 0·48 (95% CI 0·23-0·99), p=0·049 (Table 3). Neither age nor sex predicted likelihood of carriage in this study in univariate analysis and no statistically significant interaction was detected between HIV and age (p=0.481) nor sex (p=0.937) for predicting likelihood of carriage (Supplementary Figure E3). In view of the large age and sex differences between groups, the model was adjusted for age and sex resulting in an adjusted OR of 0·39 (95% CI 0·16-0·91), p=0·033 (Table 3).

**Table 2.** Experimental carriage rate by day and dose. Experimental serotype 6B pneumococcal carriage in PLHIV and HIV-uninfected participants at each inoculum dose and on each day following pneumococcal inoculation

|  | PLHIV |  | HIV-uninfected |  |
| --- | --- | --- | --- | --- |
|  | n experimental carriage positive/total inoculated |  | n experimental carriage positive/total inoculated |  |
|  | Single inoculation | Double inoculation | Single inoculation | Double inoculation |
| **At any time-point:** |  |  |  |  |
| 20 000 cfu/naris | 0/7 | .. | 1/7 | .. |
| 80 000 cfu/naris | 3/16 | 0/7 | 2/16 | 0/8 |
| 160 000 cfu/naris | 2/16 | 1/6 | 1/16 | 2/8 |
| 320 000 cfu/naris | 6/36 | 4/24 | 13/36 | 8/19 |
| **All doses** | **11/75 (15%)** | **5/37 (14%)** | **17/75 (23%)** | **10/35 (29%)** |
|  | **16/75 (21%)** |  | **27/75 (36%)** |  |
| **Day 2 post-inoculation:** |  |  |  |  |
| 20 000 cfu/naris | 0/7 | .. | 0/7 | .. |
| 80 000 cfu/naris | 1/16 | 0/7 | 1/16 | 0/8 |
| 160 000 cfu/naris | 2/16 | 1/6 | 1/16 | 1/8 |
| 320 000 cfu/naris | 2/36 | 2/22 | 10/36 | 5/18 |
| **All doses** | **5/75 (6.7%)** | **3/36 (8.3%)** | **12/75 (16%)** | **6/34 (18%)** |
| **Day 7 post-inoculation:** |  |  |  |  |
| 20 000 cfu/naris | 0/7 | .. | 1/7 | .. |
| 80 000 cfu/naris | 3/16 | 0/7 | 1/16 | 0/8 |
| 160 000 cfu/naris | 1/16 | 0/6 | 0/16 | 1/8 |
| 320 000 cfu/naris | 5/36 | 2/23 | 10/36 | 5/19 |
| **All doses** | **9/75 (12%)** | **2/36 (5.5%)** | **12/75 (16%)** | **6/35 (17%)** |
| **Day 14 post-inoculation:** |  |  |  |  |
| 20 000 cfu/naris | 0/7 | .. | 0/6* | .. |
| 80 000 cfu/naris | 2/16 | 0/7 | 0/16 | 0/8 |
| 160 000 cfu/naris | 1/16 | 0/6 | 0/16 | 1/8 |
| 320 000 cfu/naris | 4/36 | 3/23 | 7/36 | 2/17 |
| **All doses** | **7/75 (9.3%)** | **3/36 (8.3%)** | **7/75 (9.3%)** | **3/35 (8.6%)** |

Number of experimental pneumococcal carriage events defined as the presence of serotype 6B pneumococcus in nasal wash samples by classical culture on each day and at each inoculation dose. Denominator is total number who received single- or double-inoculation respectively. Entire cohort received single inoculation and subset receiving a second inoculation in a later phase of the study. Dose escalation occurred if no safety concerns had been identified at the preceding dose and experimental carriage rate in HIV-uninfected had not reached 50% as per pre-defined study protocol.
cfu=colony-forming unit
\*n=1 participant with missing data on days 14 and 21 could not attend these study visits. Participant was experimental carriage positive on day 7 and was advised by telephone to take clearance antibiotics. Included in analysis as an experimental carriage positive individual.

**Table 3:**
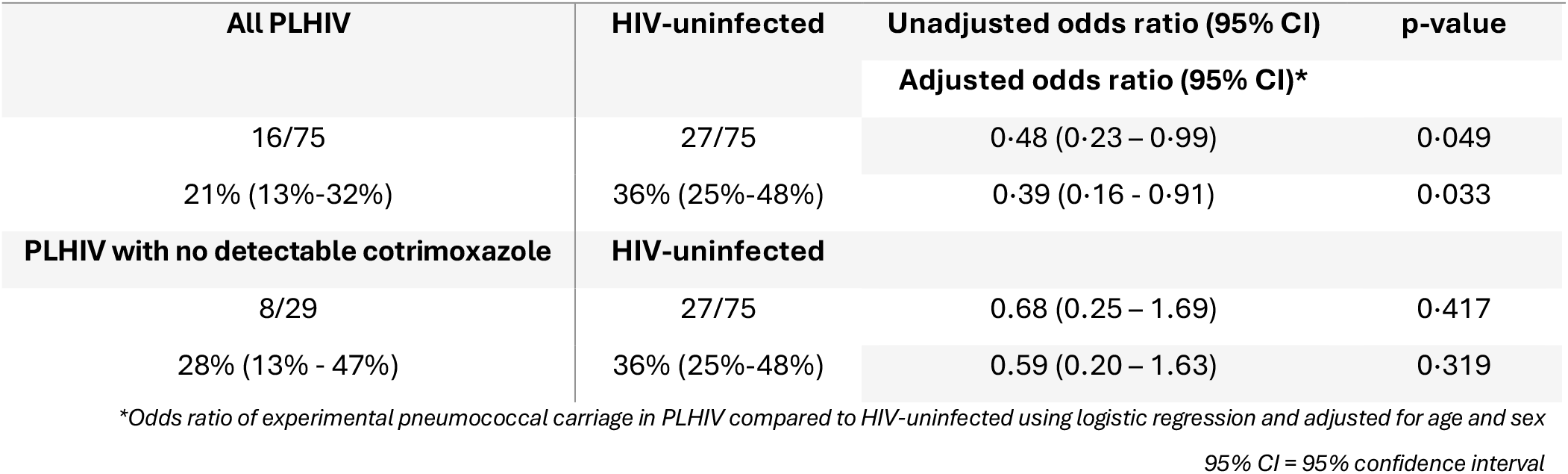
Odds of carriage in PLHIV compared to HIV-uninfected. Comparison of carriage rate in PLHIV and HIV-uninfected adults

| All PLHIV | HIV-uninfected | Unadjusted odds ratio (95% CI) | p-value |
| --- | --- | --- | --- |
|  |  | Adjusted odds ratio (95% CI)* |  |
| 16/75 | 27/75 | 0.48 (0.23 – 0.99) | 0.049 |
| 21% (13%-32%) | 36% (25%-48%) | 0.39 (0.16 - 0.91) | 0.033 |
| PLHIV with no detectable cotrimoxazole | HIV-uninfected |  |  |
| 8/29 | 27/75 | 0.68 (0.25 – 1.69) | 0.417 |
| 28% (13% - 47%) | 36% (25%-48%) | 0.59 (0.20 – 1.63) | 0.319 |

\*Odds ratio of experimental pneumococcal carriage in PLHIV compared to HIV-uninfected using logistic regression and adjusted for age and sex
95% CI = 95% confidence interval

**Figure 3.**
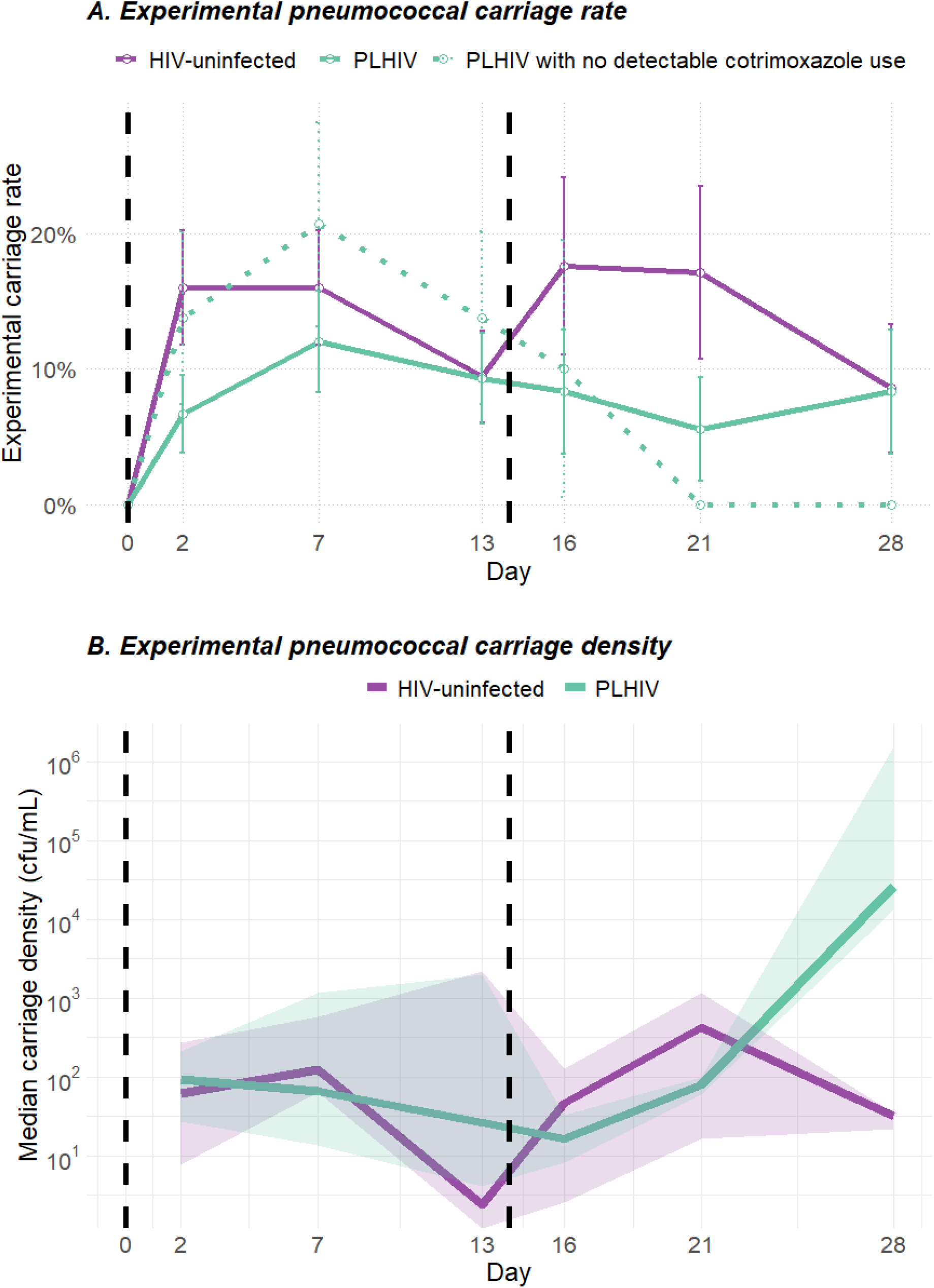
Experimental pneumococcal serotype 6B carriage in people living with HIV and HIV-uninfected. Experimental pneumococcal serotype 6B carriage in people living with HIV compared to HIV-uninfected controls. (A) Proportion of all PLHIV (turquoise solid line), HIV-uninfected (purple solid line), and PLHIV with no detectable cotrimoxazole in serum at baseline (turquoise dotted line) with experimental pneumococcal serotype 6B carriage determined by culture and latex agglutination of nasal wash samples on days 0, 2, 7, 13, 16, 21, and 28 (number with pneumococcal serotype 6B carriage/total number inoculated). Error bars represent 95% confidence intervals. Vertical dotted black lines represent pneumococcal inoculations on days 0 and 14. (B) Carriage density in 6B positive samples determined by number of colony-forming units in one millilitre of nasal wash collected from PLHIV (turquoise solid line) and HIV-uninfected (purple solid line) experimentally challenged with pneumococcus serotype 6B. Vertical dotted black lines represent pneumococcal inoculations on days 0 and 14. Solid turquoise and purple lines represent median value of 6B positive samples on that day and shaded areas represents interquartile range.

HIV-uninfected participants exhibited a dose-dependent relationship in experimental carriage rate from 14% (1/7, 95%CI 0·4%-58%) at 20000 cfu/naris, 13% (2/16, 95%CI 1·6%-38%) at 80000 cfu/naris, 19% (3/16, 95%CI 4·0%-46%) at 160000 cfu/naris, and 58% (10/21, 95%CI 41%-74%) at 320000 cfu/naris (p=0·004). This dose-dependent relationship was not detected to statistical significance in PLHIV who nonetheless showed a dose-dependent trend: 0% (0/7, 95%CI 0·0%-41%) at 20000 cfu/naris, 19% (3/16, 95%CI 4·0%-46%) at 80000 cfu/naris, 19% (3/16, 95%CI 4·0%-46%) at 160000 cfu/naris, and 28% (4/21, 95%CI 14%-45%) at 320000 cfu/naris (p=0·113) (Table 2).

Natural pneumococcal carriage rate at baseline did not differ between the groups and was not associated with rate of experimental pneumococcal carriage in PLHIV. It was associated with less experimental pneumococcal 6B carriage in HIV-uninfected participants. HIV-uninfected with natural carriage at baseline experienced experimental carriage in 22% of participants (5/23, 95% CI 7·5%-44%) compared to 42% with no natural carriage (22/52, 95% CI 29%-57%). PLHIV with natural pneumococcal carriage at baseline experienced experimental carriage in 24% of participants (6/25, 95% CI 9·4%-45%) compared to 20% without natural pneumococcal carriage at baseline (10/50, 95% CI 10%-34%) (Supplementary Table E7).

The use of molecular diagnostics for pneumococcal carriage detection in a subset of participants resulted in a higher experimental carriage rate in both groups (12/60, 20% [95% CI 11%-32%] in PLHIV and 18/55, 33% [95%CI 21%-47%] in HIV-uninfected) and also demonstrated lower carriage rate in PLHIV than HIV-uninfected controls (adjusted OR 0·38, 95% CI 0·13-1·05, p=0·067) (Supplementary table E8, Supplementary Figure E4).

### Cotrimoxazole use partially explains lower than expected experimental pneumococcal carriage rate in people living with HIV in Malawi

Cotrimoxazole was measured in baseline serum in a subset of PLHIV and was detectable in 50% (29/58). Experimental pneumococcal carriage was present in one participant (1/29, 3·4% [95% CI 0·1%-18%]) with detectable cotrimoxazole at baseline, and eight (8/29, 28% [95% CI 13%-47%], p=0·025) with no detectable cotrimoxazole at baseline (Table 3 and Figure 3A). The one participant with detectable cotrimoxazole and experimental carriage had the lowest detected concentration of the combination antibiotic (Supplementary Table E9). The adjusted odds of experimental carriage in PLHIV with no detectable cotrimoxazole compared to HIV-uninfected was lower although the results did not reach statistical significance (OR 0·59 [95% CI 0·20-1·63], p=0·319) (Table 3).

### Experimental pneumococcal serotype 6B carriage density does not differ between people living with HIV and HIV-uninfected

No statistically significant difference in overall experimental carriage density was detected between carriage positive PLHIV and HIV-uninfected by measuring the area under the curve (AUC) of carriage density over day 2 to 14 after first inoculation (median AUC 381 in PLHIV [IQR 199-11862] compared to 881 in HIV-uninfected [IQR 478-19063], p=0·547), and over day 2 to 14 after second inoculation (day 16 to 28 overall) (median AUC 348 in PLHIV [IQR 41·3-93033] compared to 189 in HIV-uninfected [IQR 18·3-5884], p=0·357) (Figure 3B).

### Experimental pneumococcal serotype 6B carriage clearance slower in people living with HIV than HIV-uninfected

HIV-uninfected with experimental carriage showed reduction of carriage rate and density between day 7 and day 14 post inoculation (for both first and second inoculation) while rate and density persist in PLHIV with experimental carriage (Table 2 and Figure 3B). By day 14 after first inoculation, 59% (10/17) of carriage positive HIV-uninfected had cleared carriage and 36% of PLHIV had done so (4/11). By day 14 after second inoculation (day 28 overall), 80% (8/10) of carriage positive HIV-uninfected had cleared carriage compared to 40% (2/5) of carriage positive PLHIV. An interval-censored Weibull survival model estimated a lower likelihood of clearance in PLHIV compared to HIV-uninfected over follow-up: Hazard ratio (HR) for clearance of carriage over days 2 to 14 after first inoculation 0·44 (95% CI 0·14-1·42) and over days 16 to 28 (after second inoculation) 0·40 (95% CI 0·08-1·93). Proportional hazards assumption tested by survival curves (Supplementary Figure E5).

## Discussion

This study establishes the first controlled human infection model (CHIM) to be conducted in PLHIV and demonstrates that these experimental systems can be conducted safely and acceptably in this potentially vulnerable population. No serious adverse events were experienced and 99% of participants would recommend participation. The rate of experimental pneumococcal carriage acquisition with serotype 6B was lower in PLHIV (21%) compared to HIV-uninfected (36%) in contrast to community studies reporting higher carriage prevalence in PLHIV.^1,5,16–18^ Exploratory analyses indicate that carriage duration may be an important factor driving the carriage burden in PLHIV, in keeping with a modelling study of community data.^1^

Cotrimoxazole use in PLHIV partially explains the lower-than-expected experimental carriage rate in PLHIV in this study. The absence of cotrimoxazole in serum of PLHIV at baseline was associated with a higher point estimate of experimental pneumococcal carriage (28%) than PLHIV without detectable cotrimoxazole in serum at baseline (3·4%), although carriage rate still did not reach that in HIV-uninfected (36%). Absence of detectable antibiotic indicates that individuals have not taken the combination antibiotic in at least the preceding two to three days but may exclude individuals who had taken cotrimoxazole prior to this and may vary according rate of renal excretion.^19^ As a tissue-penetrating antibiotic the absence in serum may not reflect absence in body tissues such as the nasal mucosa in individuals with incomplete adherence to daily cotrimoxazole. This study advocates for a more nuanced evaluation of antibiotic use when conducting CHIMs in our setting, with detection methods in tissue and blood valuable for more effective stratification of experimentally challenged populations.

The unexpected lower carriage rate in PLHIV could not be attributed to the age or sex differences between groups, nor to differences in natural pneumococcal carriage at baseline. Unmeasured sociodemographic factors, such as number of social contacts and household composition, may be a driver of high community point prevalence observed in PLHIV but were not captured in this study.^20^ Strict inclusion criteria to select PLHIV deemed safe to inoculate may have inadvertently controlled for sociodemographic drivers of high community carriage prevalence in HIV. PLHIV in this study had to demonstrate documentary evidence of ART adherence and good health for at least two years preceding study entry which may result in a participant cohort with greater health literacy and healthcare engagement than the average PLHIV in Blantyre, Malawi. Indeed, baseline natural carriage rates observed in this study differed from a community estimate in the same context, lending support to this interpretation. Natural carriage rates of 7·7% and 34% for vaccine- and non-vaccine-serotypes were observed in PLHIV in Blantyre, Malawi over a similar time period, compared to 13% and 20% in this study.^1^ HIV-uninfected participants in this study exhibited a baseline natural carriage rate of 31% compared to 25% and 27% observed previously.^1,21^ The study population excluded participants with prior pneumococcal vaccination and those living with children under five years old who are the main drivers of household and community transmission, possibly altering baseline susceptibility to carriage in the study population compared to the wider population.^22,23^

This was a single-strain infection model which used pneumococcal serotype 6B for its low invasive propensity to ensure safety of participants. A single-strain infection model cannot detect differences in carriage dynamics and immunological control across pneumococcal serotypes. As many immune mechanisms by which the pathogen invades, and the host prevents and controls pneumococcal carriage is conserved across serotypes, a single serotype models still generates relevant and vital data.^24^ The study was not designed to look at carriage duration beyond 14 days but exploratory analysis has suggested carriage duration is an important factor in the pneumococcal carriage burden observed in PLHIV. While confidence intervals were wide, the consistent hazard estimate for clearance after first and second inoculation lends weight to the finding and is in keeping with modelling data.^1^ Individuals with symptoms or recent evidence of a respiratory tract infection were excluded from this study. Asymptomatic viral or bacterial colonisation was not evaluated in this study but is an important determinant of pneumococcal carriage susceptibility and may differ by HIV status.^25,26^

## Conclusion

The EHPC in PLHIV has safely and acceptably been established and suggests prolonged carriage duration rather than increased acquisition could explain the high pneumococcal carriage burden in HIV. The carriage findings highlight the added value of a CHIM with a clear exposure time-point to progress understanding of the pneumococcal carriage burden in PLHIV. If validated these findings have important implication for pneumococcal vaccine policy in PLHIV in sub-Saharan Africa.

## Data Availability

Data from this study is available on request from the authors. 

## List of Abbreviations

95% CI: 95% confidence interval
AUC: area-under-the-curve
CHIM: controlled human infection model
EHPC: experimental human pneumococcal carriage
IQR: interquartile range
PLHIV: people living with HIV

## Acknowledgements

We are indebted to the study participants. This study would not have been possible without the support of the Data Safety Monitoring Board who ensured participant safety throughout. The DSMB was led by Professor Robert Read (University of Southampton, UK) and included Professors Johnstone Kumwenda (Kamuzu University of Health Sciences, Malawi), Neil French (University of Liverpool, UK), and Mavuto Mukaka (Mahidol-Oxford Tropical Medicine Research Unit, Thailand). We are grateful for the insightful comments on an earlier draft of this manuscript provided by Professor Homer L. Twigg III, M.D. (Indiana University Medical Centre) and Professor Robert Read (University of Southampton). This study would not have been possible without the support of local clinical services specifically Queen Elizabeth Central Hospital and Mwaiwathu Private Hospital for agreeing to facilitate inpatient care should it have been required, and Blantyre District Health Office clinics for supporting study recruitment. We are grateful to the Clinical Research Support Unit (CRSU) at Malawi-Liverpool Wellcome research programme for their expert support and guidance throughout the study.

## Contributors

SG, KJ, KD, AC conceived and designed the study with advice from HM, MH, DF, PB, JP, JR, TH. AC and KD led the study. VN, EN, EM, TH recruited participants, ensured participant safety, and collected clinical data with supervision from SG, PB, AC, KD, GL. BG, GK, CM, MPK, LC, GT, TN processed samples and generated microbiology data with supervision from TC, KJ, and JP. LM managed data. JN, AN, MA, SM advised on and generated drug level data. NT oversaw governance. KD and AC analysed the data with support from SG, KJ, DF, MH, JP, JR. KD wrote the first draft of the report; and all authors reviewed the final manuscript.

## Ethics

Ethical approval was granted by the Malawian National Health Science Research Committee (NHSRC) (22/09/3053) and by Liverpool School of Tropical Medicine’s Research Ethics Committee (22-077).

## Availability of data and materials

Data from this study is available on request from the authors.

