## Supplementary materials for "Experimental human pneumococcal carriage in adults with HIV in Malawi"

### Supplementary methods

#### Detailed pneumococcal challenge procedure

Preparation of the challenge inoculum Streptococcus pneumoniae serotype 6B strain BHN418 has been described previously.[1] Briefly, a pre-specified dose of live bacterium was prepared in sterile saline. A volume of 0·1 mL was instilled into each nostril with participants in a semi-recumbent position, taking care not to disrupt the nasal epithelium. Inoculation dose was escalated sequentially in successive cohorts from 20 000 to 80 000, 160 000, and 320 000 colony-forming units per nostril according to predefined safety and experimental carriage criteria.[2]

#### Nasal wash sampling and microbiological culture

Nasal wash samples were collected by instilling 5 mL of sterile saline twice per naris. Participants were instructed to retain the saline briefly in the nares before allowing it to drain into a sterile Galli pot. Nasal wash samples were pooled, cleared of mucus, and centrifuged to obtain a pellet. The pellet was resuspended in skim milk, tryptone, glucose, and glycerol (STGG) medium and divided into three aliquots. One aliquot was plated onto Columbia sheep blood agar (Oxoid, Basingstoke, UK) and incubated overnight at 37°C in 5% carbon dioxide. Pneumococcal colonies were identified by morphology and confirmed by optochin sensitivity and bile solubility testing. Bacterial density was determined using a dilution method.[1] Pneumococcal serotype was confirmed by latex agglutination (Immulex Pneumotest, Statens Serum Institute, Copenhagen, Denmark).

#### Molecular detection of pneumococcal carriage

Genomic bacterial DNA was extracted from a second STGG-suspended pellet using an Agowa Mag mini-DNA extraction kit (LGC Genomics, Berlin, Germany). Multiplex quantitative polymerase chain reaction was performed to identify Streptococcus pneumoniae using the lytA gene (forward primer 5′-ACGCAATCTAGCAGATGAAGCA-3′, reverse primer 5′-TCGTGCGTTTTAATTCCAGCT-3′, probe 5′-TGCCGAAAACGCTTGATACAGGGAG-3′) and to identify serotype 6B using the cpsA gene (forward primer 5′-AAGTTTGCACTAGAGTATGGGAAGGT-3′, reverse primer 5′-ACATTATGTCCATGTCTTCGATACAAG-3′, probe 5′-TGTTCTGCCCTGAGCAACTGG-3′). Reactions were run on a QuantStudio 7 Flex system and analysed using Design and Analysis software version 2.8.0 (Applied Bioscience, ThermoFisher). Experimental pneumococcal carriage was defined as detection of both lytA and cpsA gene targets in any post-inoculation sample.

#### Safety monitoring and adverse event reporting

Participants were housed in open study accommodation with a member of the study team for the first three days following inoculation for safety monitoring. Participants were provided with a safety pack containing written instructions, a thermometer, and an antibiotic course, and were instructed to record daily temperatures and report these to the study team. Participants were able to contact a study team member by telephone 24/7 and were provided with phone credit. During scheduled study visits, adverse events were actively solicited and vital signs measured by a clinical study team member. Any abnormalities prompted review by a study doctor. Serious adverse events were defined as death, hospitalisation, life-threatening events, or permanent disability. An independent Data and Safety Monitoring Board received monthly safety reports throughout recruitment. Carriage-positive participants were advised to take a three-day course of amoxicillin at study completion.

#### Acceptability assessment

Acceptability was assessed using a self-completed exit questionnaire employing a five-point Likert scale (strongly disagree, disagree, neutral, agree, strongly agree) in response to 13 statements addressing recruitment and consent, study procedures, study design and safety, study accommodation, and overall participation (Supplementary Table E3). Any negative responses prompted discussion with a study team member to explore participant feedback.

#### Additional variables

Participant age, sex, and clinical history were obtained by questionnaire and confirmed through medical record review. Serum cotrimoxazole concentrations (sulfamethoxazole and trimethoprim) were measured at the United Kingdom Antimicrobial Reference Laboratory. Any detectable drug concentration (lower limit of detection 5·0 mg/L for sulfamethoxazole and 1·0 mg/L for trimethoprim) was considered evidence of cotrimoxazole prophylaxis.

#### Statistical analysis

Statistical analyses were prespecified in the published study protocol.[2] Continuous variables were compared using Wilcoxon rank-sum tests and categorical variables using Fisher's exact tests. Confidence intervals for proportions were calculated using the Clopper–Pearson exact method. Logistic regression models were used to examine associations between participant characteristics and experimental carriage, with Wald's method used to derive confidence intervals. Dose–response relationships were assessed by logistic regression of log-transformed inoculation dose against probability of carriage. Clearance rate among experimentally colonised participants was compared separately after the first and second inoculations using interval-censored Weibull proportional hazards models, with clearance assumed to occur between the last carriage-positive and first subsequent carriage-negative visit. All analyses were two-sided and performed using R version 4.3.0.

#### Supplementary table E1: Post-inoculation study visit proforma

| Question | Response |
|---|---|
| Enter today's date | |
| Enter participants ID | |
| Enter follow up visit | Visit 3 (day 2 post-inoculation) |
| Has verbal consent been obtained to proceed? | Yes<br>No |
| HEALTH-CHECK | |
| Participant health today | Well<br>Unwell |
| Has the participant commenced antibiotics since previous review? | Yes<br>No |
| Date antibiotics commenced | |
| Reason for starting antibiotics | |
| Person who instigated starting antibiotic course | Study doctor<br>Other study team member<br>Participant themselves<br>Other |
| Does participant have any of the following symptoms? | |
| Sore throat | Yes<br>No |
| Congested or discharging nose | Yes<br>No |
| Earache | Yes<br>No |
| Feeling feverish | Yes<br>No |
| A cough | Yes<br>No |
| Sputum production | Yes<br>No |
| Headache | Yes<br>No |
| Fatigue | Yes<br>No |
| Shortness of breath | Yes<br>No |
| Chest pain | Yes<br>No |
| Joint and/or muscle pains | Yes<br>No |
| Vomiting or diarrhoea | Yes<br>No |
| Any other symptoms | Yes - please specify<br>No |
| Participant vital signs today | Tolerated values: |
| Heart rate | 60 to 90 |
| Respiratory rate | 12 to 18 |
| Oxygen saturations | >94% |
| Temperature | 36.1-37.5 |
| Systolic blood pressure | 110-180 |
| Has the participant been advised to commence antibiotics? | Yes<br>No |
| SAMPLE COLLECTION:<br>- Nasosorption<br>- Nasal wash<br>- Nasal curettage (1 and 2)<br>- Throat swab<br>- Saliva<br>- Urine<br>- Venepuncture (1x blood RNA) | |
| Nasosorption performed | Yes<br>No |
| Details of person obtaining sample | |

| | |
|---|---|
| Has the sample been labelled with participant ID | Yes<br>No |
| Nasal wash performed | Yes<br>No |
| Details of person obtaining sample | |
| Has the sample been labelled with participant ID | Yes<br>No |
| Volume of nasal wash acquired | |
| 55.1. Nasal curettage performed 1 | Yes<br>No |
| Details of person obtaining sample | |
| Has the sample been labelled with participant ID | Yes<br>No |
| Is there any nasal trauma evident following nasal curettage? | Yes<br>No |
| 55.2. Nasal curettage performed 2 | Yes<br>No |
| Details of person obtaining sample | |
| Has the sample been labelled with participant ID | Yes<br>No |
| Is there any nasal trauma evident following nasal curettage? | Yes<br>No |
| Throat swab performed | Yes<br>No |
| Details of person obtaining sample | |
| Has the sample been labelled with participant ID | Yes<br>No |
| Saliva collection performed | Yes<br>No |
| Details of person obtaining sample | |
| Has the sample been labelled with participant ID | Yes<br>No |
| Venepuncture perfomed | Yes<br>No |
| Details of person obtaining sample | |
| Has the sample been labelled with participant ID | Yes<br>No |
| 2x serum sample taken | Yes<br>No |
| 2x lithium heparin samples taken | Yes<br>No |
| 2.5 mL blood RNA sample taken | Yes |
| 60. Urine sample collected | Yes<br>No |
| Details of person obtaining sample | |
| Has the sample been labelled with participant ID | Yes<br>No |

### Supplementary table E2: CONSORT checklist

| CONSORT item | Checklist item | Section / location in manuscript |
|---|---|---|
| **Title and abstract** | | |
| 1a | Identification as a randomised trial in the title | Not applicable (non-randomised experimental human challenge study) |
| 1b | Structured summary of trial design, methods, results, and conclusions | Abstract (page 2) |
| **Introduction** | | |
| 2a | Scientific background and explanation of rationale | Introduction (page 3) |
| 2b | Specific objectives or hypotheses | Introduction (page 3) |
| **Methods** | | |
| 3a | Description of trial design | Materials and methods: Study design (page 4) |
| 3b | Important changes to methods after trial commencement (with reasons) | Materials and methods: Study design (page 4) |
| 4a | Eligibility criteria for participants | Materials and methods: Study population and setting (page 4) |
| 4b | Settings and locations where the data were collected | Materials and methods: Study population and setting (page 4) |
| 5 | Interventions for each group with sufficient detail to allow replication | Materials and methods: Pneumococcal challenge and safety monitoring (pages 4–5); Supplementary Methods |
| 6a | Completely defined pre-specified primary and secondary outcome measures | Material and methods: Outcome measure: Experimental pneumococcal carriage (page 5); Supplementary Methods |
| 6b | Any changes to trial outcomes after the trial commenced (with reasons) | NA |
| 7a | How sample size was determined | Materials and methods: Sample size and statistical analysis (page 5) |
| 7b | When applicable, explanation of interim analyses and stopping guidelines | Materials and methods: Pneumococcal challenge and safety monitoring (pages 4–5); Supplementary Methods |
| **Randomisation and blinding** | | |
| 8a–10 | Sequence generation, allocation concealment, implementation | NA |
| 11a–11b | Blinding | NA |
| **Statistical methods** | | |
| 12a | Statistical methods used to compare groups for primary and secondary outcomes | Materials and methods: Sample size and statistical analysis (page 5); Supplementary Materials |
| 12b | Methods for additional analyses (e.g. adjusted and subgroup analyses) | Supplementary Methods |
| **Results** | | |
| 13a | Participant flow (numbers enrolled, inoculated, followed up, analysed) | Supplementary Figure E2 |
| 13b | Losses and exclusions after enrolment (with reasons) | Supplementary Figure E2 |
| 14a | Dates defining recruitment and follow-up | Materials and methods: Study population and setting (page 4) |
| 14b | Why the trial ended or was stopped | Materials and methods: Sample size and statistical analysis (page 5); Supplementary Materials |
| 15 | Baseline demographic and clinical characteristics | Results: Comparison groups and sociodemographic characteristics (page 6); Table 1 |
| 16 | Number of participants included in each analysis | Results (pages 6); Table 1 |
| 17a | Outcomes and estimation (effect sizes and precision) | Results (pages 6-9); Tables 2–3; Figures 2–3 |
| 17b | For binary outcomes, presentation of absolute and relative effect sizes | Results (pages 6-9); Tables 2–3; Figures 2–3 |

| | | |
|---|---|---|
| 18 | Results of any other analyses performed | Results; Supplementary Tables E7–E9; Supplementary Figures E3–E4 |
| **Harms** | | |
| 19 | All important harms or unintended effects in each group | Results: Safety and acceptability (pages 6–7); Figure 2; Supplementary Tables E5–E6 |
| **Discussion** | | |
| 20 | Trial limitations, addressing sources of potential bias | Discussion (pages 10–11) |
| 21 | Generalisability (external validity) | Discussion (pages 10–11) |
| 22 | Interpretation consistent with results, balancing benefits and harms | Discussion (pages 10–11) |
| **Other information** | | |
| 23 | Registration number and name of trial registry | Study design, Materials and methods (page 4) |
| 24 | Where the full trial protocol can be accessed | Peer-reviewed publication; ClinicalTrials.gov (NCT05698225) |
| 25 | Sources of funding and other support | Abstract (page 3); Funding statement (page 12) |

#### Supplementary figure E1: Study design

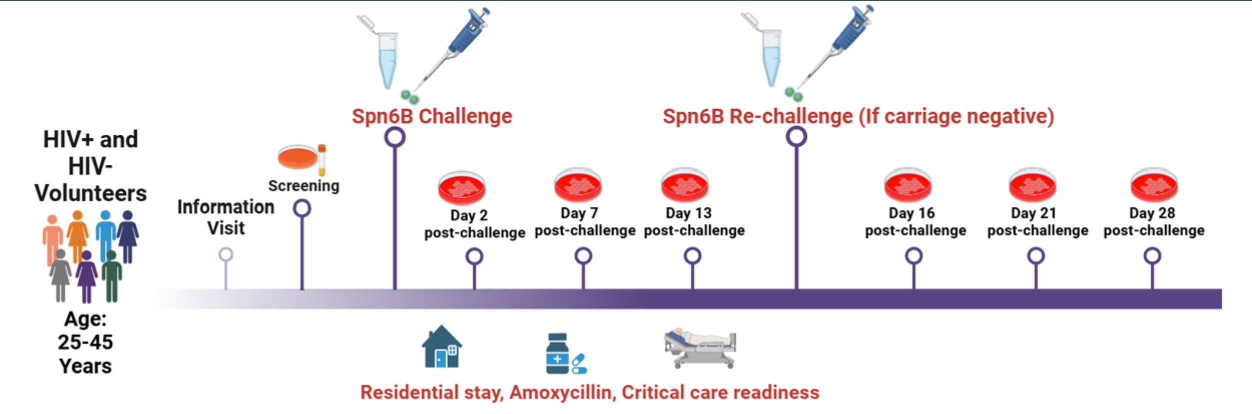

*Supplementary figure E1: Study design. Potential participants attended for an information visit which included verbal and written information in English and Chichewa. After a cooling-off period, if potential participants then wanted to participate, they would have to self-present to the study clinic rooms to complete formal screening including a screening questionnaire with a clinical study team member, a review of medical records, a clinical examination, and screening blood tests. Eligible participants had a pre-specified dose of Streptococcus pneumoniae serotype 6B introduced into both nostrils using a pipette. Participants resided in study accommodation with a member of the study team for three days following inoculation. Participants attended for in-person study visits on days two, seven, and 13 or 14 following first inoculation to identify any symptoms or clinical signs and to take nasal and blood samples. The presence of Streptococcus pneumoniae 6B in the nose at a post-inoculation study visit was defined as experimental pneumococcal carriage and was determined by classical culture of nasal wash samples. After safety of one inoculation was established, a second phase of the study included a second inoculation on day 14 in participants who were carriage negative on days two, seven, and 13. Participants were re-inoculated with the same dose of bacteria and then sampled to identify experimental pneumococcal carriage on days 16, 21, and 28. Participants were issued with a safety pack which included a three-day course of amoxicillin, written study and safety information, contact details for the study doctor on call 24/7, and a thermometer. The study had agreements with a local private and public hospital on how to treat any participants requiring inpatient care however this was not required during the study.*

### Supplementary results tables

#### Supplementary table E3: Acceptability of model to participants

**Supplementary table E3: Acceptability of the experimental carriage model:** Participants self-completed the acceptability questionnaire on their final study visit using a Likert scale to evaluate each statement against strongly agree, agree, neutral, disagree, or strongly disagree. Number and percentages reflect participants who responded with strongly agree or agree.

| | PLHIV<br>n (%) who agree with the statements<br>n=74 | HIV-uninfected<br>n (%) who agree with the statements<br>n=73 |
|---|---|---|
| **1. Recruitment and consent** | | |
| 1b. The way I heard about this study was appropriate and did not interfere with my clinical care | 72 (97%) | 72 (99%) |
| 1c. I received sufficient information about the study before consenting to participate | 73 (99%) | 70 (96%) |
| 1d. I was given sufficient time to consider the study before consenting to participate | 73 (99%) | 71 (97%) |
| **2. Study procedures** | | |
| 2a. The procedure to have the bacteria placed in my nose did not cause undue discomfort | 70 (95%) | 72 (99%) |
| 2b. The procedures to take samples from my nose did not cause undue discomfort | 69 (93%) | 67 (92%) |
| **3. Study design and safety** | | |
| 3a. The frequency of visits during the study was appropriate | 73 (99%) | 71 (97%) |
| 3b. I felt the steps taken to keep me safe during the study were appropriate (for example; daily text messages with study team, health-checks during study visits, study accommodation) | 72 (97%) | 71 (97%) |
| 3c. I was able to contact the study team easily for advice | 72 (97%) | 73 (100%) |
| **4. Study accommodation** | | |
| 4a. The accommodation provided after inoculation was satisfactory | 71 (96%) | 73 (100%) |
| 4b. I felt 3 days was an appropriate time to remain in study accommodation | 71 (96%) | 69 (95%) |
| **5. Study participation** | | |
| 5a. The clinical team treated me with respect and kindness throughout the study | 74 (100%) | 73 (100%) |
| 5b. The compensation provided by the study was appropriate | 67 (91%) | 67 (92%) |
| 5c. I would recommend participation in this study to a friend | 73 (99%) | 72 (99%) |

### Supplementary table E4: Natural carriage serotypes at baseline

| Supplementary table E4: Natural carriage serotypes at baseline | | |
|---|---|---|
| | **PLHIV** | **HIV-uninfected** |
| **All serotypes** | **25/75 (33%)** | **23/75 (32%)** |
| **Vaccine serotype** | **10 (13%)** | **6 (8.0%)** |
| 14 | 1 | 0 |
| 19 | 2 | 2 |
| 23 | 2 | 2 |
| 3 | 2 | 0 |
| 6A | 0 | 1 |
| 7 | 1 | 1 |
| 9 | 2 | 0 |
| **Non-vaccine serotypes** | **15 (20%)** | **17 (23%)** |
| 33 | 0 | 1 |
| 8 | 0 | 2 |
| 10 | 2 | 1 |
| 15 | 1 | 2 |
| 20 | 0 | 1 |
| NVT | 12 | 10 |

### Supplementary table E5: Number of adverse events in each group

| Supplementary table E5: Safety of experimental pneumococcal serotype 6B challenge in PLHIV and HIV-uninfected participants | | | |
|---|---|---|---|
| | **PWH** | **HIV-uninfected** | |
| Serious adverse event* | 0 | 0 | |
| Adverse events | 19% (n=14/75) | 13% (n=10/75) | p=0·505 |
| Sore throat | 2 | 2 | |
| Cough | 3 | 6 | |
| with sputum | 0 | 1 | |
| Coryzal symptoms | 1 | 3 | |
| Feverish | 1 | 1 | |
| Headache | 5 | 4 | |
| Fatigue | 0 | 0 | |
| Earache | 0 | 0 | |
| Shortness of breath | 0 | 0 | |
| Fever | 1 | 0 | |
| Vomiting | 1 | 0 | |
| Diarrhoea | 0 | 1 | |
| Joint pains | 2 | 0 | |
| Chest pain | 0 | 1 | |
| Other symptoms** | 5 | 1 | |

*Serious adverse event defined as death, hospitalisation, life-threatening event, or permanent disability
** n=3 abdominal pain, n=1 unknown, n=1 eyelid itchiness, n=1 rash

### Supplementary table E6: Adverse events by experimental carriage status

| Supplementary table E6: Adverse events by carriage status | | | |
|---|---|---|---|
| **Adverse event** | **Carriage negative (n=107)** | **Carriage positive (n=43)** | p-value |
| Any adverse event | 17 (16%) | 7 (16%) | 0.953 |
| Cough | 5 (5%) | 4 (9%) | 0.28 |
| Coryzal symptoms | 3 (3%) | 1 (4%) | 1 |
| Headache | 7 (7%) | 2 (5%) | 0.659 |
| Sore throat | 3 (3%) | 1 (2%) | 1 |
| Feverish | 1 (1%) | 1 (2%) | 0.388 |

#### Supplementary table E7: Experimental carriage rate by natural carriage at baseline

| Supplementary table E7: Experimental carriage rate by natural carriage status at baseline | | | | |
|---|---|---|---|---|
| | **PWH** | | **HIV-uninfected** | |
| | **Natural carriage at baseline** | **No natural carriage** | **Natural carriage at baseline** | **No natural carriage** |
| **Experimental carriage rate**<br>Proportion (95% CI) | 6/25<br>24% (9.4% - 45%) | 10/50<br>20% (10% - 34%) | 5/23<br>22% (7.5% - 44%) | 22/52<br>42% (29% - 57%) |

#### Supplementary table E8: Comparison of experimental carriage by PCR-based methods in PLHIV and HIV-uninfected

| Supplementary table E8: Comparison of carriage rate by PCR (95% confidence interval) in PLHIV and HIV-uninfected adults | | | |
|---|---|---|---|
| **All PLHIV** | **HIV-uninfected** | **OR (95% CI)**<br>**aOR* (95% CI)** | **p-value** |
| 12/60<br>20% (11%-32%) | 18/55**<br>33% (21%-47%) | 0.51 (0.22-1.19) | 0.123 |
| | | 0.38 (0.13-1.05) | 0.0672 |
| **PLHIV with no detectable cotrimoxazole** | **HIV-uninfected** | | |
| 8/29<br>28% (13%-47%) | 18/55*<br>33% (21%-47%) | 0.78 (0.28-2.07) | 0.628 |
| | | 0.65 (0.19-2.00) | 0.460 |

**adjusted for age and sex, **n=5 participants removed due to serotype 6B carriage detected by molecular methods at baseline*

#### Supplementary table E9: Concentration of sulfamethoxazole and trimethoprim in people living with HIV with detectable cotrimoxazole at baseline

| | Sulfamethoxazole concentration (mg/L) | Trimethoprim concentration (mg/L) | Experimental Carriage |
|---|---|---|---|
| 1. | 11.0 | <1.0 | No |
| 2. | 12.2 | <1.0 | No |
| 3. | 17.1 | <1.0 | No |
| 4. | 20.2 | <1.0 | No |
| 5. | 27.5 | <1.0 | No |
| 6. | 28.9 | 1.1 | No |
| 7. | 29.3 | <1.0 | No |
| 8. | 29.4 | 2.0 | No |
| 9. | 29.5 | <1.0 | No |
| 10. | 33.5 | 1.4 | No |
| 11. | 36.9 | <1.0 | No |
| 12. | 38.8 | 2.9 | No |
| 13. | 40.2 | 1.6 | No |
| 14. | 41.1 | <1.0 | No |
| 15. | 46.2 | 1.5 | No |
| 16. | 48.6 | <1.0 | No |
| 17. | 51.5 | 1.3 | No |
| 18. | 55.5 | 1.8 | No |
| 19. | 58.3 | 1.9 | No |
| 20. | 58.3 | 1.5 | No |
| 21. | 62.6 | 1.3 | No |
| 22. | 68.8 | 1.7 | No |
| 23. | 7.5 | <1.0 | Yes |
| 24. | 72.0 | 1.9 | No |
| 25. | 74.5 | 2.1 | No |
| 26. | 74.5 | 1.9 | No |
| 27. | 76.4 | 2.5 | No |
| 28. | 80.2 | 1.9 | No |
| 29. | 90.0 | 2.6 | No |

### Supplementary results figures

#### Supplementary figure E2: Study population and inoculation protocols

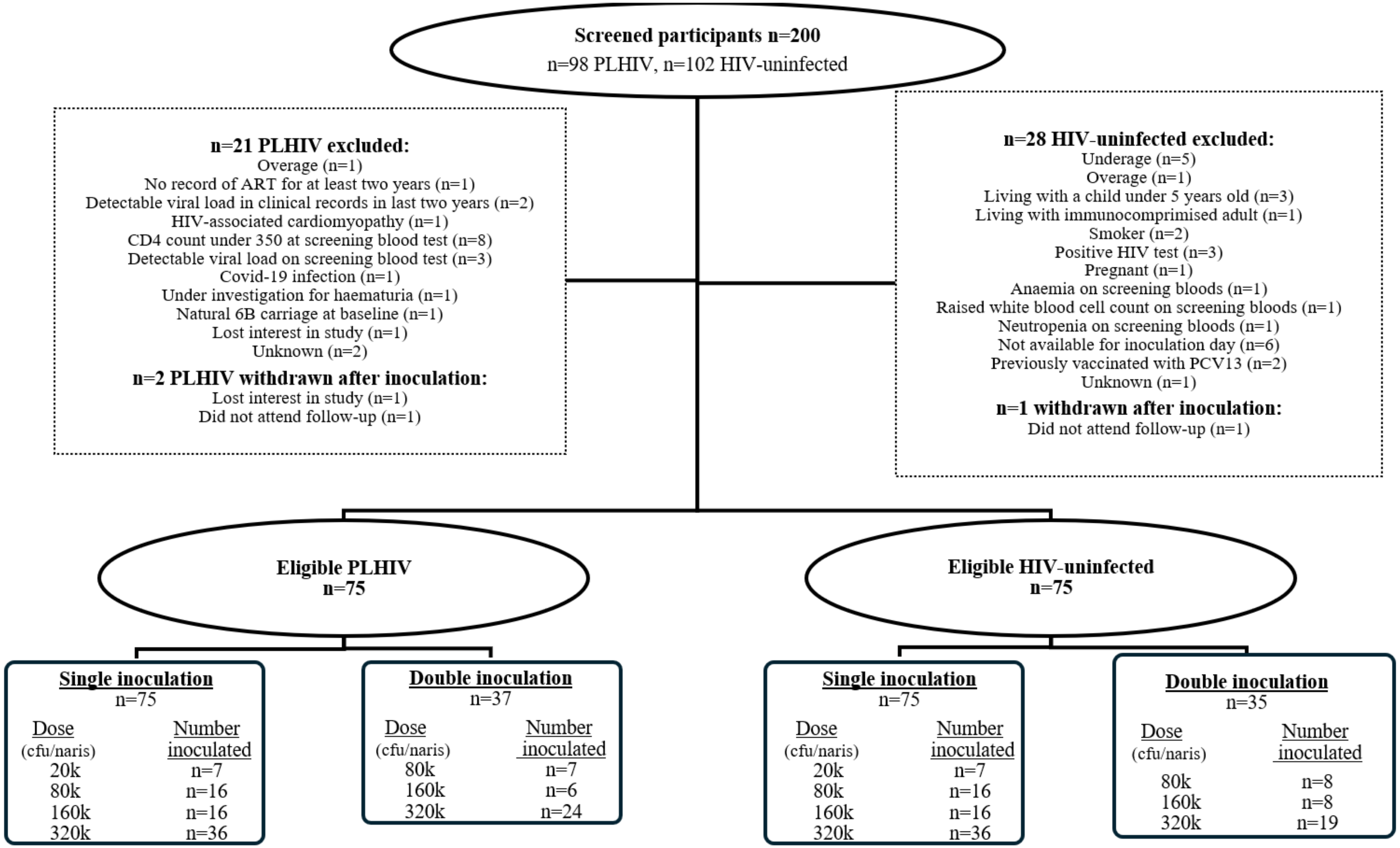

*Supplementary figure E2: Potential participants were screened after self-presenting to study clinic rooms if they were able to demonstrate eligibility criteria (e.g. proof of age, medical records from preceding two years) before completing screening questionnaire with a member of the study team, clinical examination, and bloods tests to determine eligibility. Of 200 screened participants, 77 PLHIV and 76 HIV-uninfected controls were deemed eligible for inoculation. Two PLHIV and one HIV-uninfected participant were withdrawn after inoculation. All participants received one inoculation and in a second study phase (after safety with one inoculation had been demonstrated) all carriage negative participants received a second inoculation on day 14. The pre-specified dose-escalation protocol stipulated that at each dose a review of safety and carriage rate should be completed and if carriage rate was below 50% and no safety issues were identified, the study should proceed to the next dose. cfu=colony forming unit, PLHIV=people living with HIV*

#### Supplementary figure E3: Odds of Experimental Carriage by HIV status, age, and sex.

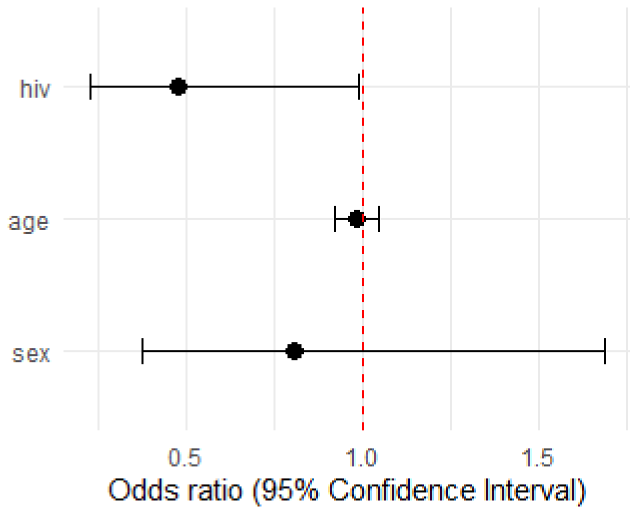

#### Supplementary figure E4: Carriage rate by PCR over time

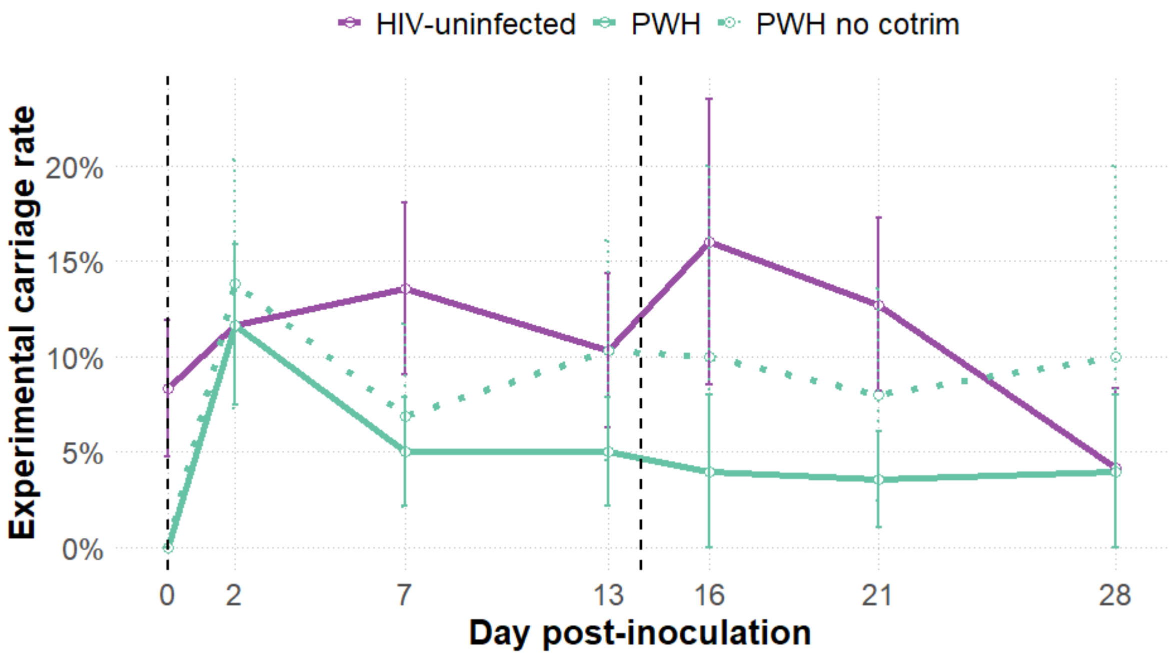

#### Supplementary figure E5: Carriage clearance over follow-up

**A**

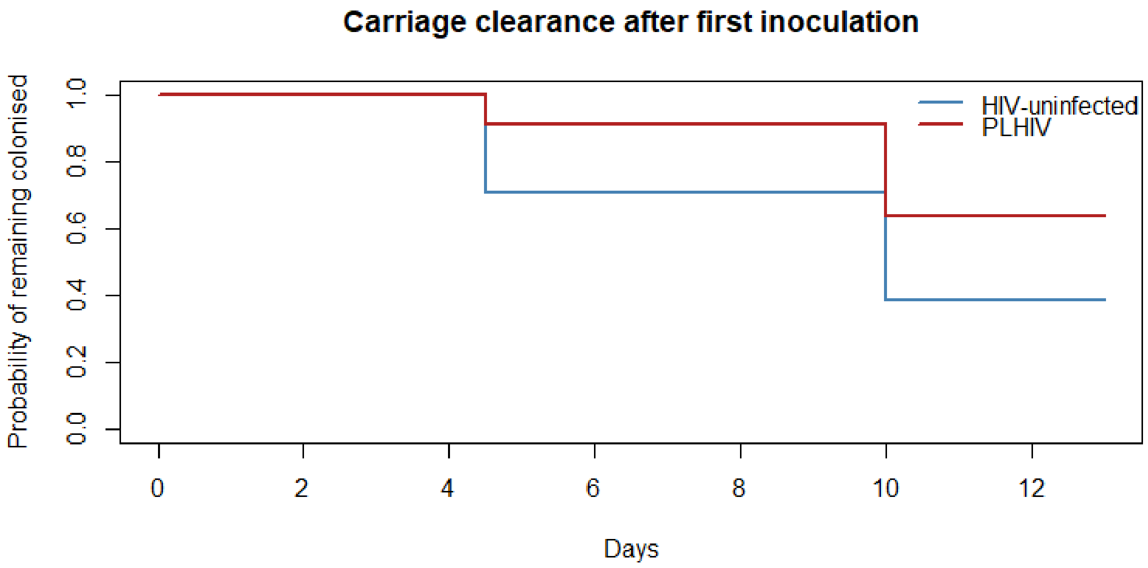

**B**

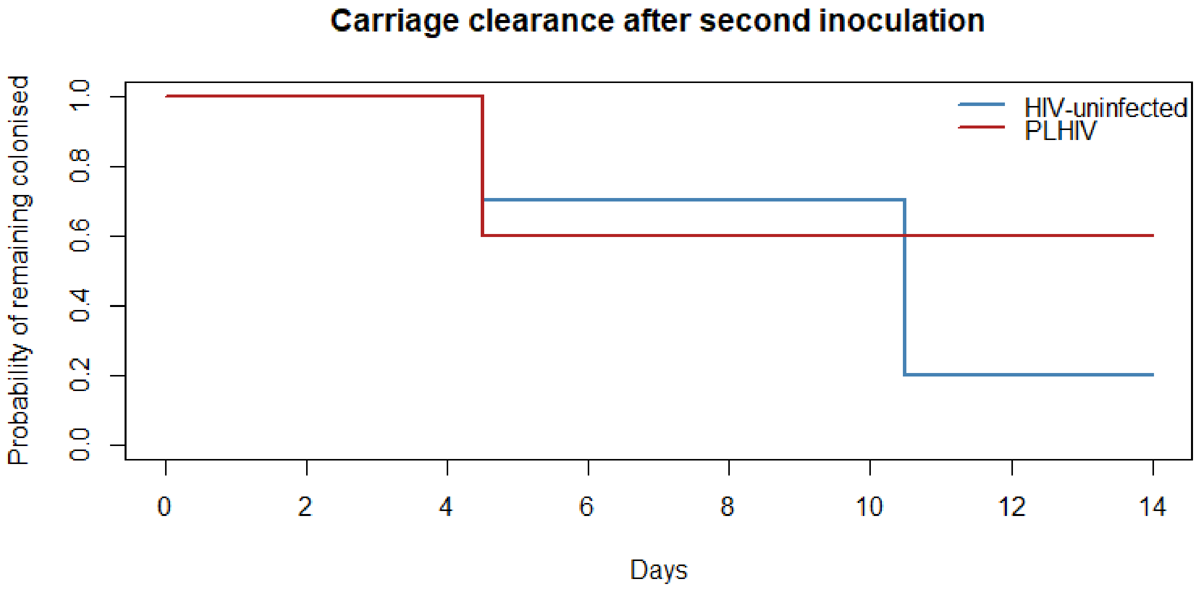

*Supplementary Figure E5: Nonparametric interval-censored (Turnbull) survival curves showing the probability of remaining colonised with serotype 6B pneumococcus over time following (A) first inoculation (days 0–14) and (B) second inoculation (days 16–28). Analyses include only participants who developed experimental carriage during the respective inoculation period. Clearance was defined as the first negative culture following a positive sample, with the exact time of clearance interval-censored between study visits (days 2, 7, and 14 after inoculation).*
